## Supplementary material for "Clinical manifestations and disease severity of SARS-CoV-2 infection among infants in Canada"

**eFigure 1. Odds ratios* of COVID-19 admission by continuous infant age, compared to infants at 100 days of age.**

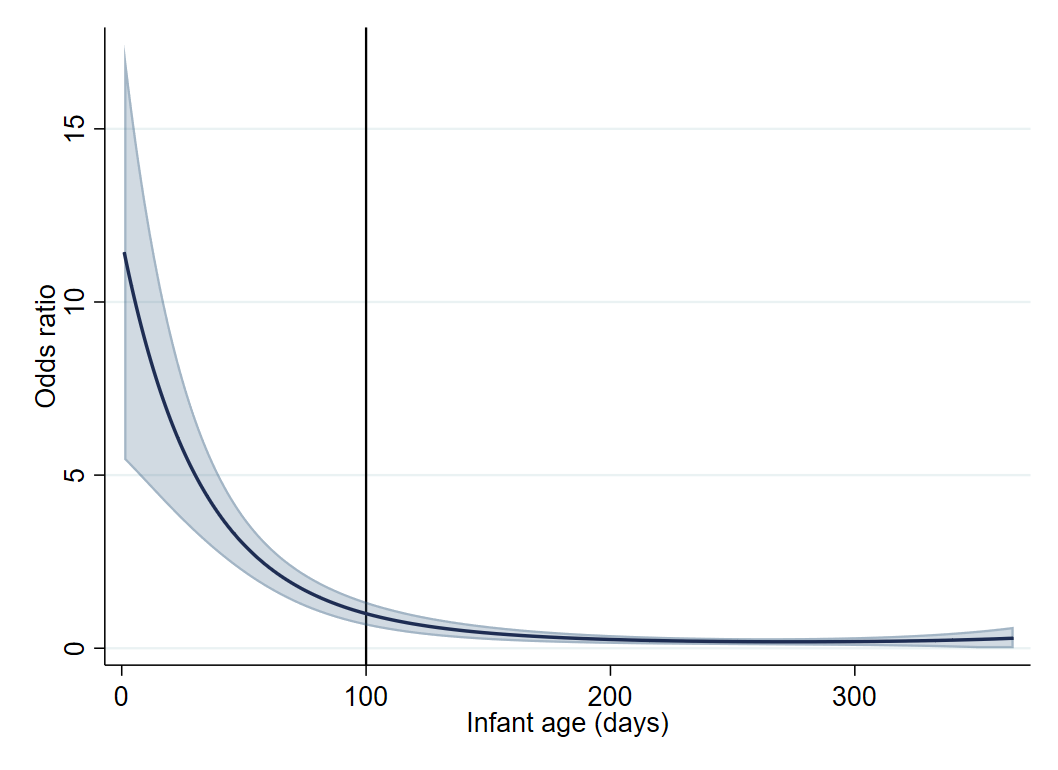

*ORs are adjusted by infant sex, gestational age category, comorbid conditions, and wave of pandemic. Continuous infant age was analyzed as a quadratic term.

**eTable 1. SARS-CoV-2 CPSP Severity criteria**

| **Severity category** | **Clinical criteria** | **Outcome criteria** |
| --- | --- | --- |
| **Asymptomatic** | 4.2: No symptoms reported, or write-in note specifying “asymptomatic”  4.4: No clinical features reported / no symptoms of COVID-19  OR  6.1, 6.2, 6.3: All presenting symptoms explained by concurrent infection | 7.1: No targeted treatment against COVID-19 received  7.4: No respiratory support required  AND  Patient does not have any of the criteria for mild, moderate or severe disease. |
| **Outpatient care** | 4.2: Any reported symptoms of COVID-19  4.4: Clinical syndromes may include gastrointestinal symptoms, skin changes, bronchiolitis or pneumonia | 4.1. No admission required  7.1: No targeted treatment against COVID-19  7.3: Managed at home  7.4: No respiratory support required  AND  Patient does not have any of the criteria for moderate or severe disease. |
| **Mild disease**  **(in-patient)** | 4.2: Reported symptoms may include fever, cough, sore throat, coryza, sneezing, lethargy (only for patients ≥1 month/≥1 year old), skin manifestations, muscle aches, rash, vomiting, diarrhea, loss of appetite, conjunctivitis, headache, loss of smell, lethargy, or loss of taste.  4.4: Clinical syndromes may include gastrointestinal symptoms, skin changes, bronchiolitis or pneumonia | 4.1. Admission required  7.1: No targeted treatment against COVID-19  7.3: Inpatient ward  7.4: No respiratory support required  AND  Patient does not have any of the criteria for moderate or severe disease. |
| **Severity category** | **Clinical criteria** | **Outcome criteria** |
| **Moderate disease** | Any of the presenting symptoms described in the mild category | 7.1: Targeted treatment against COVID-19, including Remdesivir, steroids, hydroxychloroquine/chloroquine, anti-IL1 or anti-IL6  7.4: Increased baseline home oxygen or low-flow oxygen  AND  Patient does not have any of the criteria for severe disease. |
| **Severe disease (any one of the listed criteria)** | 4.2: Seizures, coma  4.4: Acute respiratory distress syndrome (ARDS), cytokine storm, seizures, stroke, encephalitis, encephalopathy, acute necrotizing encephalopathy, coma, hypotension, acute cardiac dysfunction | 7.3: Intensive care unit admission  7.4: High-flow nasal cannula. Non-invasive ventilation (e.g., CPAP or BiPAP), Conventional mechanical ventilation, High-frequency oscillatory ventilation, Nitric oxide (NO), Extracorporeal membrane oxygenation (ECMO) or vasopressors.  7.6. Death (secondary to COVID-19) |

**eTable 2. Logistic regression sensitivity analysis of factors associated with hospital admission due to COVID-19-related disease.**

| **Characteristics** | | Odds ratio for COVID-19 hospitalization (hospitalizations unrelated to COVID-19 grouped with outpatient cases) | | | | Odds ratio for COVID-19 hospitalization (restricted to all CHUSJ, HSC, and MCH cases^1^) | | | |
| --- | --- | --- | --- | --- | --- | --- | --- | --- | --- |
|  |  | Crude model | | Adjusted model^2^ | | Crude model | | Adjusted model^3^ | |
|  |  | OR (95% CI) | *P* value | aOR (95% CI) | *P* value | OR (95% CI) | *P* value | aOR (95% CI) | *P* value |
| **Infant age** | |  |  |  |  |  |  |  |  |
|  | 0–<1 month | 2.60 (1.49–4.52) | 0.001 | 2.90 (1.62–5.17) | <0.001 | 3.89 (1.75–8.65) | 0.001 | 3.41 (1.49–7.80) | 0.004 |
|  | 1–3 months | 1 [Reference] | NA | 1 [Reference] | NA | 1 [Reference] | NA | 1 [Reference] | NA |
|  | 4–6 months | 0.32 (0.18–0.59) | <0.001 | 0.29 (0.15–0.54) | <0.001 | 0.42 (0.18–0.94) | 0.035 | 0.30 (0.12–0.74) | 0.009 |
|  | 7–12 months | 0.18 (0.10–0.32) | <0.001 | 0.15 (0.08–0.29) | <0.001 | 0.25 (0.11–0.54) | <0.001 | 0.18 (0.07–0.42) | <0.001 |
| **Infant sex** | |  |  |  |  |  |  |  |  |
|  | Female | 1 [Reference] | NA | 1 [Reference] | NA | 1 [Reference] | NA | 1 [Reference] | NA |
|  | Male | 1.20 (0.81–1.77) | 0.36 | 1.02 (0.65–1.59) | 0.93 | 1.81 (1.03–3.20) | 0.04 | 1.74 (0.91–3.33) | 0.09 |
| **Gestational age at birth** | |  |  |  |  |  |  |  |  |
|  | Term (≥37 weeks') | 1 [Reference] | NA | 1 [Reference] | NA | 1 [Reference] | NA | 1 [Reference] | NA |
|  | Late preterm (34–<37 weeks') | 1.02 (0.44–2.36) | 0.96 | 0.72 (0.28–1.87) | 0.50 | 0.98 (0.27–3.58) | 0.97 | 0.98 (0.22–4.27) | 0.98 |
|  | Moderate/very preterm (<34 weeks') | 1.58 (0.70–3.54) | 0.27 | 1.28 (0.51–3.20) | 0.60 | 1.96 (0.57–6.72) | 0.29 | 2.21 (0.55–8.95) | 0.27 |
| **Comorbid conditions** | |  |  |  |  |  |  |  |  |
|  | None/Unknown | 1 [Reference] | NA | 1 [Reference] | NA | 1 [Reference] | NA | 1 [Reference] | NA |
|  | ≥1 comorbid condition | 1.25 (0.69–2.25) | 0.47 | 2.70 (1.32–5.53) | 0.007 | 2.28 (1.01–5.13) | 0.047 | 4.89 (1.79–13.35) | 0.002 |
| **Phase of COVID-19 pandemic** | |  |  |  |  |  |  |  |  |
|  | 1st wave (April–August 2020) | 1 [Reference] | NA | 1 [Reference] | NA | 1 [Reference] | NA | 1 [Reference] | NA |
|  | 2nd wave (September 2020–February 2021) | 1.26 (0.70–2.27) | 0.44 | 1.48 (0.77–2.85) | 0.24 | 0.68 (0.33–1.42) | 0.30 | 0.76 (0.33–1.73) | 0.51 |
|  | 3rd wave (March–May 2021) | 1.27 (0.68–2.38) | 0.45 | 1.89 (0.93–3.82) | 0.08 | 0.62 (0.28–1.36) | 0.23 | 0.72 (0.29–1.80) | 0.49 |
| aOR = Adjusted odds ratio; OR = Odds ratio | | | | | | | | | |
| ^1^Centre Hospitalier Universitaire Sainte-Justine (Montreal), Hospital for Sick Children (Toronto), and Montreal Children's Hospital (Montreal). | | | | | | | | | |
| ^2^Multivariable analysis was conducted among 495 complete cases. | | | | | | | | | |
| ^3^Multivariable analysis was conducted among 317 complete cases. | | | | | | | | | |

**eTable 3. Characteristics of infants with SARS-CoV-2 infection reported from the Centre Hospitalier Universitaire Sainte-Justine, Hospital for Sick Children, or Montreal Children's Hospital.**

| **Characteristics** | | | All Infants | Outpatients | Inpatients | | *P* value |
| --- | --- | --- | --- | --- | --- | --- | --- |
|  |  |  |  |  | Not COVID-19 related | COVID-19 related |  |
| **Total cases reported, N** | | | 374 | 275 | 33 | 66 | --- |
| **Infant age (days), median (IQR)** | | | 142 (52–253) | 160 (73–263) | 139 (52–262) | 37 (22–115) | <0.001 |
| **Infant age category, n (%)** | | |  |  |  |  | <0.001 |
|  | <1 month | | 44 (11.8) | 16 (5.8) | 6 (18.2) | 22 (33.3) | --- |
|  | 1–3 months | | 97 (25.9) | 65 (23.6) | 9 (27.3) | 23 (34.8) | --- |
|  | 4–6 months | | 84 (22.5) | 68 (24.7) | 6 (18.2) | 10 (15.2) | --- |
|  | 7–12 months | | 149 (39.8) | 126 (45.8) | 12 (36.4) | 11 (16.7) | --- |
| **Infant sex, n (%)** | | |  |  |  |  | 0.11 |
|  | Female | | 160 (42.8) | 126 (45.8) | 13 (39.4) | 21 (31.8) | --- |
|  | Male | | 214 (57.2) | 149 (54.2) | 20 (60.6) | 45 (68.2) | --- |
| **Population group of infant, n (%)^1^** | | |  |  |  |  | --- |
|  | White | | 36 (9.6) | 21 (7.6) | 5 (15.2) | 10 (15.2) | 0.08 |
|  | South Asian | | 43 (11.5) | 27 (9.8) | 5 (15.2) | 11 (16.7) | 0.21 |
|  | Arab/West Asian | | 26 (7.0) | 10–13 (3.6–4.7) | <5 (<15.2) | 12–15 (18.2–22.7) | <0.001 |
|  | Black | | 23 (6.1) | 12 (4.4) | 5 (15.2) | 6 (9.1) | 0.03 |
|  | East/Southeast Asian | | 14 (3.7) | 10 (3.6) | <5 (<15.2) | <5 (<7.6) | 0.66 |
|  | Indigenous | | <5 (<1.3) | <5 (<1.8) | 0 (0.0) | 0 (0.0) | --- |
|  | Other | | 11 (2.9) | 7–10 (2.5–3.6) | <5 (<15.2) | 0 (0.0) | >0.99 |
|  | Unknown | | 228 (61.0) | 188 (68.4) | 13 (39.4) | 27 (40.9) | <0.001 |
| **Gestational age at birth, n (%)^2^** | | |  |  |  |  | <0.001 |
|  | Term (≥37 weeks') | | 312 (89.1) | 231 (92.0) | 22 (66.7) | 59 (89.4) | --- |
|  | Late preterm (34–<37 weeks') | | 22 (6.3) | 11–14 (4.4–5.6) | 6–9 (18.2–27.3) | <5 (<7.6) | --- |
|  | Moderate/very preterm (<34 weeks') | | 16 (4.6) | 8 (3.2) | <5 (<15.2) | <5 (<7.6) | --- |
|  |  | Median (IQR) weeks at birth^3^ | 34.6 (31.9–35.7) | 34.4 (32.3–35.9) | 34.6 (27.0–36.0) | 31.9 (29.0–35.6) | 0.62 |
| **Any comorbid condition, n (%)** | | | 44 (11.8) | 20 (7.3) | 14 (42.4) | 10 (15.2) | <0.001 |
| **Any co-infections, n (%)** | | | 26 (7.0) | 16 (5.8) | 5 (15.2) | 5 (7.6) | <0.001 |
| **COVID-19 exposure, n (%)** | | |  |  |  |  | --- |
|  | Known close contact with confirmed SARS-CoV-2 infection | | 211 (56.4) | 146 (53.1) | 14 (42.4) | 51 (77.3) | <0.001 |
|  | Nosocomial infection | | <5 (<1.3) | 0 (0.0) | <5 (<15.2) | 0 (0.0) | <0.001 |
| **SARS-CoV-2 lineage, n (%)** | | |  |  |  |  |  |
|  | Cases before December 26, 2020^4^ | | 145 (38.8) | 102 (37.1) | 15 (45.4) | 28 (42.4) | --- |
|  | Cases occurring after December 26, 2020 | | 229 (61.2) | 173 (62.9) | 18 (54.6) | 38 (57.6) | --- |
|  |  | Alpha (B.1.1.7) | 39 (17.0) | 31 (17.9) | <5 (<27.8) | 5–8 (13.2–21.1) | --- |
|  |  | VOC, other or unspecified^5^ | 21 (9.2) | 17 (9.8) | <5 (<27.8) | <5 (<13.2) | --- |
|  |  | VOC not detected | 35 (15.3) | 29 (16.8) | 0 (0.0) | 6 (15.8) | --- |
|  |  | VOC screening not conducted | 134 (58.5) | 96 (55.5) | 15 (83.3) | 23 (60.5) | --- |
| ^1^Physicians could report multiple population groups. East/Southeast Asian includes Chinese, Filipino, Japanese, Korean, and Southeast Asian. Indigenous includes First Nations, Inuit, and Métis. | | | | | | | |
| ^2^Gestational age category not known for 24 outpatients. | | | | | | | |
| ^3^Among preterm-born infants only (i.e. <37 weeks' gestation). | | | | | | | |
| ^4^i.e. Date of first Alpha detection in Canada. | | | | | | | |
| ^5^Includes B1351, P1, and variants with N501Y mutation but otherwise unspecified. | | | | | | | |

**eTable 4. Characteristics of admitted infants with severe COVID-19**

| **Characteristics, n (%)** | | Frequency | Percent |
| --- | --- | --- | --- |
| **Admissions with severe COVID-19, N** | | 20 | --- |
| **Admitted to ICU** | | 14 | 70.0 |
| **Any respiratory/hemodynamic support required** | | 15 | 75.0 |
|  | Low-flow oxygen | 7 | 35.0 |
|  | High-flow nasal cannula | 5 | 25.0 |
|  | Non-invasive ventilation | <5 | <25.0 |
|  | Mechanical ventilation^1^ | 5 | 25.0 |
|  | Vasopressors | 0 | 0.0 |
| **Clinical syndromes** | |  |  |
|  | URTI | 8 | 40.0 |
|  | Pneumonia | 5 | 25.0 |
|  | Acute cardiac dysfunction | <5 | <25.0 |
|  | Acute respiratory distress syndrome | <5 | <25.0 |
|  | Bronchiolitis | <5 | <25.0 |
|  | Encephalopathy | <5 | <25.0 |
|  | Gastrointestinal | <5 | <25.0 |
|  | Hematologic disorder^2^ | <5 | <25.0 |
|  | Seizure(s) | <5 | <25.0 |
| **Radiologic findings** | |  |  |
|  | Abnormal chest x-ray | 8 | 40.0 |
|  | Abnormal CT scan | 0 | 0.0 |
| **Infant died** | | <5 | <25.0 |
| URTI=Upper respiratory tract infection. | | | |
| ^1^Includes conventional mechanical and high-frequency oscillatory ventilation. | | | |
| ^2^Includes anemia, lymphopenia, neutropenia, and thrombocytosis. | | | |

**eTable 5. Symptoms, laboratory findings, and clinical syndromes of infants with SARS-CoV-2 infection by age group.**

| **Clinical features** | | Infant age category | | |
| --- | --- | --- | --- | --- |
|  |  | <1 month | 1–12 months | *P* value^1^ |
| **COVID-19-related hospitalizations, N^2^** | | 49 | 91 | --- |
| **COVID-19 severity, n (%)** | |  |  | 0.03 |
|  | Asymptomatic/outpatient care | --- | --- | --- |
|  | Inpatient care, mild/moderate | 37 (75.5) | 83 (91.2) | --- |
|  | Inpatient care, severe | 12 (24.5) | 8 (8.8) | --- |
| **Symptoms and signs, n (%)** | |  |  |  |
|  | Fever | 36 (73.5) | 70 (76.9) | 0.65 |
|  | Runny nose | 21 (42.9) | 35 (38.5) | --- |
|  | Decreased oral intake | 17 (34.7) | 33 (36.3) | 0.85 |
|  | Respiratory distress | 13 (26.5) | 21 (23.1) | --- |
|  | Cough | 12 (24.5) | 39 (42.9) | --- |
|  | Lethargy | 11 (22.4) | 16 (17.6) | --- |
|  | Sneezing | 8 (16.3) | 5 (5.5) | --- |
|  | Vomiting | 6 (12.2) | 20 (22.0) | --- |
|  | Diarrhea | 7 (14.3) | 16 (17.6) | --- |
|  | Irritability^3^ | 5 (10.2) | 8 (8.8) | --- |
|  | Rash | <5 (<10.2) | 6 (6.6) | --- |
|  | Conjunctivitis | <5 (<10.2) | 5 (5.5) | --- |
|  | Tachycardia^3^ | <5 (<10.2) | <5 (<5.5) | --- |
| **Laboratory findings, n (%)** | |  |  |  |
|  | Neutropenia | 5 (10.2) | 16 (17.6) | --- |
|  | Lymphopenia | <5 (<10.2) | 12 (13.2) | --- |
|  | Anemia | <5 (<10.2) | <5 (<5.5) | --- |
|  | Thrombocytosis | 0 (0.0) | 6 (6.6) | --- |
| **Clinical syndromes, n (%)** | |  |  |  |
|  | Upper respiratory tract infection | 27 (55.1) | 55 (60.4) | --- |
|  | Gastrointestinal | 14 (28.6) | 30 (33.0) | --- |
|  | Bronchiolitis | <5 (<10.2) | 12 (13.2) | --- |
|  | Pneumonia | <5 (<10.2) | 8 (8.8) | --- |
|  | Coagulopathy | <5 (<10.2) | <5 (<5.5) | --- |
|  | Seizure(s) | <5 (<10.2) | <5 (<5.5) | --- |
|  | Acute respiratory distress syndrome | <5 (<10.2) | 0 (0.0) | --- |
|  | Cardiac dysfunction | <5 (<10.2) | 0 (0.0) | --- |
|  | Hepatitis | 0 (0.0) | <5 (<5.5) | --- |
| **Abnormal CXR, n (%) / cases with imaging** | | 9/30 (30.0) | 15/55 (27.3) | 0.79 |
| **Any respiratory support required, n (%)** | | 13 (26.5) | 10 (11.0) | 0.02 |
|  | Low-flow oxygen | 8 (16.3) | 7 (7.7) | --- |
|  | High-flow nasal cannula | <5 (<10.2) | <5 (<5.5) | --- |
|  | Non-invasive ventilation | <5 (<10.2) | <5 (<5.5) | --- |
|  | Mechanical ventilation^4^ | <5 (<10.2) | <5 (<5.5) | --- |
|  | Vasopressors | 0 (0.0) | 0 (0.0) | --- |
| **Admitted to ICU, n (%)** | | 8 (16.3) | 6 (6.6) | 0.08 |
| **Infant died, n (%)** | | 0 (0.0) | <5 (<5.5) | --- |
| ^1^Statistical tests conducted only for comparisons identified *a priori*. | | | | |
| ^2^Excludes 58 hospitalizations not related to COVID-19, and one COVID-19-related hospitalization for whom an age category could not be assigned. | | | | |
| ^3^Symptoms included as write-in notes only and may therefore be underrepresented. | | | | |
| ^4^Includes conventional mechanical and high-frequency oscillatory ventilation. | | | | |

**eTable 6. Treatments administered to infants with COVID-19**

| **Characteristics, n (%)** | | Frequency | Patient group | | | Child age | | | |
| --- | --- | --- | --- | --- | --- | --- | --- | --- | --- |
|  |  |  | Outpatients | Inpatient, COVID-19-related | P value | <1 month | 1–3 months | 4–12 months | P value |
| **Total patients, N (excludes hospitalizations not related to COVID-19)^1^** | | 473 | 332 | 141 | --- | 69 | 138 | 262 | --- |
| **Prescribed any treatment** | |  |  |  | <0.001 |  |  |  | <0.001 |
|  | Yes | 98 (20.7) | 18 (5.4) | 80 (56.7) | --- | 35 (50.7) | 34 (24.6) | 29 (11.1) | --- |
|  | No | 375 (79.3) | 314 (94.6) | 61 (43.3) | --- | 34 (49.3) | 104 (75.4) | 233 (88.9) | --- |
| **Treatments received** | |  |  |  |  |  |  |  |  |
|  | Antibioitics | 94 (19.9) | 16 (4.8) | 78 (55.3) | <0.001 | 35 (50.7) | 34 (24.6) | 25 (9.5) | <0.001 |
|  | Steroids | 11 (2.3) | --- | --- | --- | --- | --- | --- | --- |
|  | Immunoglobulin (IVIG) | <5 (<1.1) | --- | --- | --- | --- | --- | --- | --- |
|  | Remdesivir | <5 (<1.1) | --- | --- | --- | --- | --- | --- | --- |
|  | Other^2^ | 6 (1.3) | --- | --- | --- | --- | --- | --- | --- |
| ^1^N=58. Age category not available for 4 infants. | | | | | | | | | |
| ^2^Includes antivirals other than remdesivir, aspirin, and bronchodilators. | | | | | | | | | |

**eTable 7. Comparison of CPSP and modified Dong severity criteria**

| **CPSP classification** | | **Modified Dong classification** | |
| --- | --- | --- | --- |
| Category | n (%) | Category | n (%) |
| Asymptomatic | 66 (12.4) | Asymptomatic | 66 (12.4) |
| Outpatient care | 310 (58.4) | Mild disease | 334 (62.9) |
| Inpatient - mild disease | 125 (23.5) | Moderate disease | 79 (14.9) |
| Inpatient - moderate disease | 10 (1.9) | Severe disease | 28 (5.3) |
| Inpatient - severe disease | 20 (3.8) | Critical disease | 24 (4.5) |
